# Effects of bilateral sequential theta-burst stimulation on functional connectivity in treatment-resistant depression: first results

**DOI:** 10.1101/2022.02.16.22271078

**Authors:** Peter Stöhrmann, Godber Mathis Godbersen, Murray Bruce Reed, Jakob Unterholzner, Manfred Klöbl, Pia Baldinger-Melich, Thomas Vanicek, Andreas Hahn, Rupert Lanzenberger, Siegfried Kasper, Georg S. Kranz

## Abstract

**Background:** Previous studies suggest that transcranial magnetic stimulation exerts antidepressant effects by altering functional connectivity (FC). However, knowledge about this mechanism is still limited. Here, we aimed to investigate the effect of bilateral sequential theta-burst stimulation (TBS) on FC in treatment-resistant depression (TRD) in a sham-controlled longitudinal study.

**Methods:** TRD patients (n = 20) underwent a three-week treatment of intermittent TBS of the left and continuous TBS of the right dorsolateral prefrontal cortex (DLPFC). Upon this trial’s premature termination, 15 patients had received active TBS and five patients sham stimulation. Resting-state functional magnetic resonance imaging was performed at baseline and after treatment. FC (left and right DLPFC) was estimated for each participant, followed by group statistics (T-tests). Furthermore, depression scores were analyzed (linear mixed models analysis) and tested for correlation with FC.

**Results:** Both groups exhibited reductions of depression scores, however, there was no significant main effect of group, or group and time. Anticorrelations between DLPFC and the subgenual cingulate cortex (sgACC) were observed for baseline FC, corresponding to changes in depression severity. Treatment did not significantly change DLPFC-sgACC connectivity, but significantly reduced FC between the left stimulation target and bilateral anterior insula.

**Conclusions:** Our data is compatible with previous reports on the relevance of anticorrelation between DLPFC and sgACC for treatment success. Furthermore, FC changes between left DLPFC and bilateral anterior insula highlight the effect of TBS on the salience network.

**Limitations:** Due to the limited sample size, results should be interpreted with caution and are of exploratory nature.

## Introduction

Bihemispheric prefrontal theta-burst stimulation (TBS) involving excitatory, intermittent TBS (iTBS) of the left, and inhibitory, continuous TBS (cTBS) of the right dorsolateral prefrontal cortex (DLPFC) is a promising treatment approach for major depressive disorder (MDD) [1]. Bilateral TBS, a type of transcranial magnetic stimulation (TMS), has shown a superior patient outcome when compared to unilateral therapy [2]. Similarly, repeated TMS (rTMS) including left high-frequency (HF) and right low-frequency (LF) stimulation was shown to be superior to unilateral left HF rTMS [3], see also [4] for a review. MDD may be better addressed by modulating both rather than only one hemisphere, as several neuropsychological and imaging studies, e.g., [5-8], support a prefrontal asymmetry hypothesis of MDD, which poses a hypoactivity of the left, and a hyperactivity of the right DLPFC [9].

Previous research has looked into TMS related effects on neural activity using functional magnetic resonance imaging (fMRI). Here, estimates of functional connectivity (FC) are used to determine the communication between brain areas [10]. Notably, studies provided convincing evidence that a negative FC (i.e., anticorrelation) between the stimulation target at the left DLPFC and the subgenual anterior cingulate cortex (sgACC) is relevant for treatment success [11-16]. It was further indicated that treatment response to a variety of antidepressant treatments including TMS is associated with distinct FC changes in cortical and subcortical regions [17].

Especially non-invasive brain stimulation including TMS is seen as crucial to normalize cognitive control networks that are believed to be altered in MDD, such as the salience network (SN) and central executive network, which encompass areas such as the DLPFC, cingulate cortex and anterior insula (AI) [18, 19]. Evidence for such an enhancement comes for example from a study on healthy subjects, showing that HF rTMS of the left DLPFC selectively increases ACC connectivity towards a meso-corticolimbic network [20]. However, an inhibition of the left DLPFC using cTBS was also shown to increase connectivity between ACC, bilateral AI and the stimulated region in healthy controls [21]. Other studies did not observe changes in cognitive control networks upon HF rTMS of the left DLPFC in MDD [22]. Instead, stimulation reduced FC between sgACC and regions of interest in the default mode network (DMN) albeit these changes were not related to antidepressant treatment response [22]. In another study, accelerated iTBS of the left DLPFC (5 daily sessions spread over 4 days) increased sgACC FC to the medial orbitofrontal cortex in responders compared to non-responders, but this effect was seemingly independent from the stimulation itself [23]. Similarly, the same authors showed that HF rTMS to the left DLPFC in MDD affects FC only in responders but not in non-responders [24], while others corroborated FC changes in responders, irrespective of whether active or sham stimulation was applied [25]. Together, these results indicate that connectivity changes are related to clinical improvement rather than the mechanism of action of the stimulation itself.

Due to the heterogeneous responses to TMS treatment reported, in the current study we aimed to further elucidate FC changes upon brain stimulation with bilateral sequential TBS in pharmacologically treatment-resistant depression (TRD). We hypothesized characteristic FC changes compared to baseline for the treatment group after 3 weeks of bilateral TBS treatment, in networks altered in mood disorders, while investigating predictors for treatment response at baseline.

## Materials and methods

### Participants

A total of 20 (12 female, 8 male) right-handed patients with TRD, who took part in a more comprehensive, multimodal neuroimaging clinical trial (ClinicalTrials.gov Identifier: NCT02810717), were included. TRD was defined as failure to respond to two adequate medication trials of at least 4 weeks each in sufficient dosage for the current depressive episode as indicated in literature [26]. Patients were eligible for inclusion if they met DSM 4 criteria for single or recurrent MDD and had a Clinical Global Impression Scale score of ≥ 4 and a Hamilton depression rating scale (HAMD-17) total score of ≥ 18. Patients had to be on stable psychopharmacological treatment within four weeks prior to inclusion and were required to maintain their original medication regimen throughout the study. Exclusion criteria were a medical history of a major systemic illness (dating back not more than five years), neurological diseases, a history of a seizure, or any contraindications to MRI or TMS as screened by safety screening questionnaires [27]. Further exclusion criteria were substance abuse or dependence within the last three months prior to inclusion, a body weight of over 115 kg, pregnancy, active suicidal intent, or intake of benzodiazepines other than Lorazepam > 2mg/d or any dose of an anticonvulsant.

The study was approved by the Ethics Committee of the Medical University of Vienna (EK 1761/2015) and performed following the Declaration of Helsinki.

### Study design and treatment protocol

The study was designed as a longitudinal, patient- and assessor-blinded, sham-controlled mono-center study. After inclusion, participants underwent a baseline MRI scan and clinical assessment. They were then randomly assigned to receive daily (Monday to Friday) active sequential bilateral TBS or sham stimulation for three weeks. Participants underwent a second MRI scan and clinical assessment within one week after the last TBS session. Follow-up assessments were performed two and four weeks after the final TBS session, respectively. After completion, participants were unblinded and the sham group was offered TBS treatment.

Daily treatments consisted of two TBS sessions, separated by one hour as previously done [2]. During each session, iTBS and cTBS was administered, with iTBS targeted to the left and cTBS targeted to the right DLPFC, respectively. The stimulation sequence, i.e. order of hemispheres treated, was reversed for every consecutive session. TBS was administered using a MagPro X100 magnetic stimulator (MagVenture, Tonica Elektronik A/S, Denmark) and a figure-of-eight shaped, liquid-cooled coil (Cool-B70), with a focality of 13.9 cm^2^ (r_1/2_ = 2.1 cm) and a stimulation (i.e. half-value) depth of d_1/2_ = 1.35 cm [28]. We followed the original TBS protocol described by Huang et al., comprising 3-pulse 50-Hz bursts, applied at 5 Hz [29]. iTBS consisted of a 2-second train of theta-bursts and an inter-train-interval of 8 seconds with 20 repeated trains, whereas cTBS consisted of a continuous train of bursts, amounting to a total number of 600 pulses for each hemisphere and a total number of 1200 pulses per session. Stimulation was delivered at an intensity of 120% of the individual resting motor threshold (RMT), determined before the first treatment using visual inspection as done previously [30].

Stimulation targets were identified using neuro-navigation (LOCALITE^®^ TMS Navigator Germany) at the initial appointment and marked on personalized head-caps for later reference. The left DLPFC target was defined at Montreal Neurological Institute (MNI) coordinate [-38, +44, +26] as done previously [11], whereas the right DLPFC target was defined contralaterally at MNI coordinate [+38, +44, +26] on the patients’ normalized anatomical scan in MNI space. Sham stimulation was performed with the coil angled 90° away from the skull as previously described [3]. This produced some scalp sensation and a sound intensity comparable to active stimulation. Throughout the study, all participants, as well as clinical and research personnel handling participants were blinded (excluding the TMS operator). Possible side effects such as headache, nausea or dizziness were assessed after each stimulation.

### Clinical assessment

The primary clinical endpoint was the change in HAMD-17 score at the follow-up assessment, two weeks after the last of 15 treatment days. Secondary endpoints included changing scores of the Beck Depression Inventory (BDI-II) and the Inventory of Depressive Symptomatology (IDS-C).

### Image acquisition

MRI scans were acquired on a 3 Tesla Siemens Magnetom Prisma system (Siemens Medical, Erlangen, Germany) with a 64-channel head-neck coil. Anatomical scans were attained with a T1-weighted sequence (TE/TR = 2.91/2000 ms, 192 slices, matrix size 240 × 256 × 192, voxel size 1.00 × 1.00 × 1.00 mm^3^). Resting-state parameters were: 2D single-shot gradient-recalled EPI, TE/TR = 30/2050 ms, interleaved slice order, matrix size 100 × 100 × 35, Series Length: 176 frames (6 min, 0.8 sec), Voxel Dimensions (X, Y, Z): 2.1 × 2.1 × 2.8 (+25 % gap) mm^3^.

### Image data processing and analysis

Functional images were processed in MATLAB R2018b (The MathWorks Inc, Natick, Massachusetts) using SPM12 v6225, custom scripts and toolboxes mentioned below. fMRI scans were corrected for interleaved, ascending acquisition (“slice time”) and motion, with one subject being excluded whose scans exhibited a frame wise displacement (FD) ≥ 0.5 in ≥ 10% of consecutive time frames. Then, brain images were spatially normalized to the MNI template, masked using MNI tissue probabilities (sum of gray matter (p_GM_), white matter (p_WM_) and cerebrospinal fluid (p_CSF_) probabilities ≥ 0.5), temporally despiked using the BrainWavelet Toolbox v2.0, [31, 32], threshold = 20, chsearch = ‘harsh’), and spatially smoothed (FWHM = 8 mm, Gaussian kernel, implicit mask: p_GM_ ≥ 0.3). Time series were further regressed with global [33] and tissue specific signals (principal components derived from mean WM and CSF signals), realignment parameters including lag (difference of one frame) and their squares (Friston-24) and were temporally filtered (0.01 - 0.1 Hz).

DLPFC stimulation regions of interest (ROIs) were defined as spheres (r = 5 mm) in MNI space. Their respective centers were individually placed in the cortex, at 1.5 times the coil’s stimulation depth below the scalp [28], additionally accounting for coil orientation as recorded by the neuro-navigation system prior to initial TMS administration. This approach ensured that ROIs were both within the cortex, and magnetic field strength was comparable between different subjects at the volumes of interest. sgACC time courses were derived by weighting whole brain signals with their connectivity towards the sgACC, resulting in less noise than if extracted directly from a small ROI, as previously done [13].

ROI mean time courses were extracted using MarsBaR [34] and correlated with the whole brain on the voxel level. The resulting connectivity maps (r-scores) of the brains were Fisher-z-transformed.

Functionally defined DLPFC targets were located by anatomically masking the DLPFC, thresholding sgACC FC maps at the lowest 10%, and taking the centroid of the most anticorrelated cluster [13].

### Statistical analysis

The primary clinical endpoint was a HAMD-17 score after 15 treatment days, analyzed using a linear mixed effects model incorporating treatment group, time, group by time interaction, and patient as a random effect. Categorical treatment response was defined as any reduction from baseline HAMD-17 > 50%, and remission was additionally defined as a HAMD-17 score < 7. Alpha was set to 0.05 and analyses were conducted in SPSS (IBM SPSS Statistics, Version 27.0. Armonk, NY: IBM Corp).

Group-level statistics (one-spample, one-sided T-tests) on imaging data were calculated using SPM12, testing for significant connectivity on the cluster level (p_uncorr_ < 0.001, p_FWE, Cluster_ < 0.05) in baseline and post-treatment scans, as well as their changes (M2-M1). Furthermore, we looked for correlations between depression scores and their changes, with DLPFC-sgACC FC throughout the course of the study, although these data were not fully available for each subject. Thus, we applied tests for the subset of subjects with complete data, as well as for all subjects with missing values at two weeks post-treatment replaced by those of four weeks post-treatment (next observation carried backwards).

All analyses were run on an exploratory basis and were not corrected for the number of seeds, contrast directions and correlations.

## Results

### Study termination and clinical outcome

The study was terminated prematurely (December 2019) due to irreparable damage of the imaging equipment. By then, 20 patients, aged 38.2 ± 12.2 years, with a full set of MRI scans matching the previously described criteria were available, including 15 patients receiving active stimulation and 5 patients allocated to the sham stimulation group. Demographic and clinical information of participants are depicted in Table 1. Overall improvement in terms of a reduction of HAMD-17 compared to baseline was observed over both groups (main effect of time with F = 21.583, p < 0.001, linear mixed models analysis) There was no significant main effect of treatment group (F = 0.307, p = 0.582) and no significant interaction between group and time (F = 1.787, p = 0.188); for mean values, see Table 1. Separate analysis of the active TBS group, yielded significant effects of time with larger effect size (F = 37.422 p < 0.001). One participant in the sham group reported an increase in depressive symptoms. In the treatment group, 8 participants (53%) and in the sham group, two participants (40%) were classified as responders. 3 participants, exclusively having received active stimulation, were classified as remitters after treatment.

**Table 1:** Demographics and clinical characteristics of study participants. Post-treatment scores were collected two and four weeks after the last TBS session, i.e. five (seven) weeks after the initial TBS treatment. The medication intake was stable prior to inclusion, and during the study, here displayed in four groups: namely drug-free (DF), intake of a selective serotonin reuptake inhibitor only (SSRI: incl. escitalopram, paroxetine), or intake of another antidepressant drug only (ATD: incl. amitryptiline, mirtazapine, melitracen, bupropione, mirtazapine, lithium, levothyroxine, opipramole), as well of combinations of substance groups (comb.), containing further substance groups, namely serotonin-noradrenaline reuptake inhibitors (SNRI: incl. venlafaxine, duloxetine, milnacipran), antipsychotic drugs (APD: incl. prothipendyl, quetiapine, flupentixole), benzodiazepines (BZD: incl. lorazepam), channel blockers (CB: incl. pregabaline, lamotrigine) or positive allosteric modulators (PAM: incl. zolpidem). For mean values and standard deviations, missing values at two weeks were replaced by the next observation (Next Observation Carried Backwards, NOCB).

|  |  | **Total** | **Treatment** | **Sham** |
| --- | --- | --- | --- | --- |
| **Age** |  | 38.2 ± 12.2 | 36.1 ± 11.6 | 44.4 ± 13.2 |
| **Sex** | F/M | 12/8 | 8/7 | 4/1 |
| **Medication** | DF/SSRI/ATD/comb. | 4/3/1/12 | 3/3/1/8 | 1/0/0/4 |
| comb.: | SSRI | 4 | 4 | 0 |
|  | SNRI | 6 | 3 | 3 |
|  | APD | 6 | 4 | 2 |
|  | BZD | 2 | 0 | 2 |
|  | CB | 5 | 3 | 2 |
|  | PAM | 1 | 0 | 1 |
|  | ATD | 9 | 6 | 3 |
| **Baseline** | **HAMD-17** | 21.7 ± 3.9 | 20.5 ± 3.0 | 25.2 ± 4.7 |
|  | **IDS-C** | 37.5 ± 8.4 | 35.4 ± 5.8 | 43.6 ± 12.3 |
|  | **BDI-II** | 33.1 ± 8.2 | 31.9 ± 7.2 | 36.6 ± 10.9 |
| **Post-Treatment**<br>(2 weeks, NOCB) | **HAMD-17** | 12.5 ± 7.2 | 10.2 ± 5.6 | 19.2 ± 7.9 |
|  | **IDS-C** | 25.0 ± 13.8 | 22.1 ± 13.0 | 33.8 ± 13.5 |
|  | **BDI-II** | 24.1 ± 13.8 | 21.4 ± 12.4 | 32.0 ± 16.2 |
|  | **Responders** | 10 (50%) | 8 (53%) | 2 (40%) |
|  | **Remitters** | 3 (15%) | 3 (20%) | 0 (0%) |
|  | **Worsened** | 1 (5%) | 0 (0%) | 1 (20%) |

### Baseline functional connectivity

To determine whether individual stimulation targets at left and right DLPFC were anticorrelated with the sgACC in the entire sample, we conducted a whole-brain voxel-wise analysis for the weighted (“Seedmap Approach”) sgACC time courses [13]. Results showed that both stimulation regions aimed for (MNI coordinates [±38, +44, +26]) coincided with clusters of voxels that are significantly anticorrelated with the sgACC on group level for all 20 subjects at a significance level of p_FWE,Cluster_ = 0.05. On the voxel level (p_FWE,Peak_ < 0.05) the left DLPFC cluster was found to be slightly inferior to the stimulation site (see Fig. 1). In line with above observations, 90% of all the stimulation sites were anticorrelated in the respective individuals’ connectivity maps.

**Figure 1:**
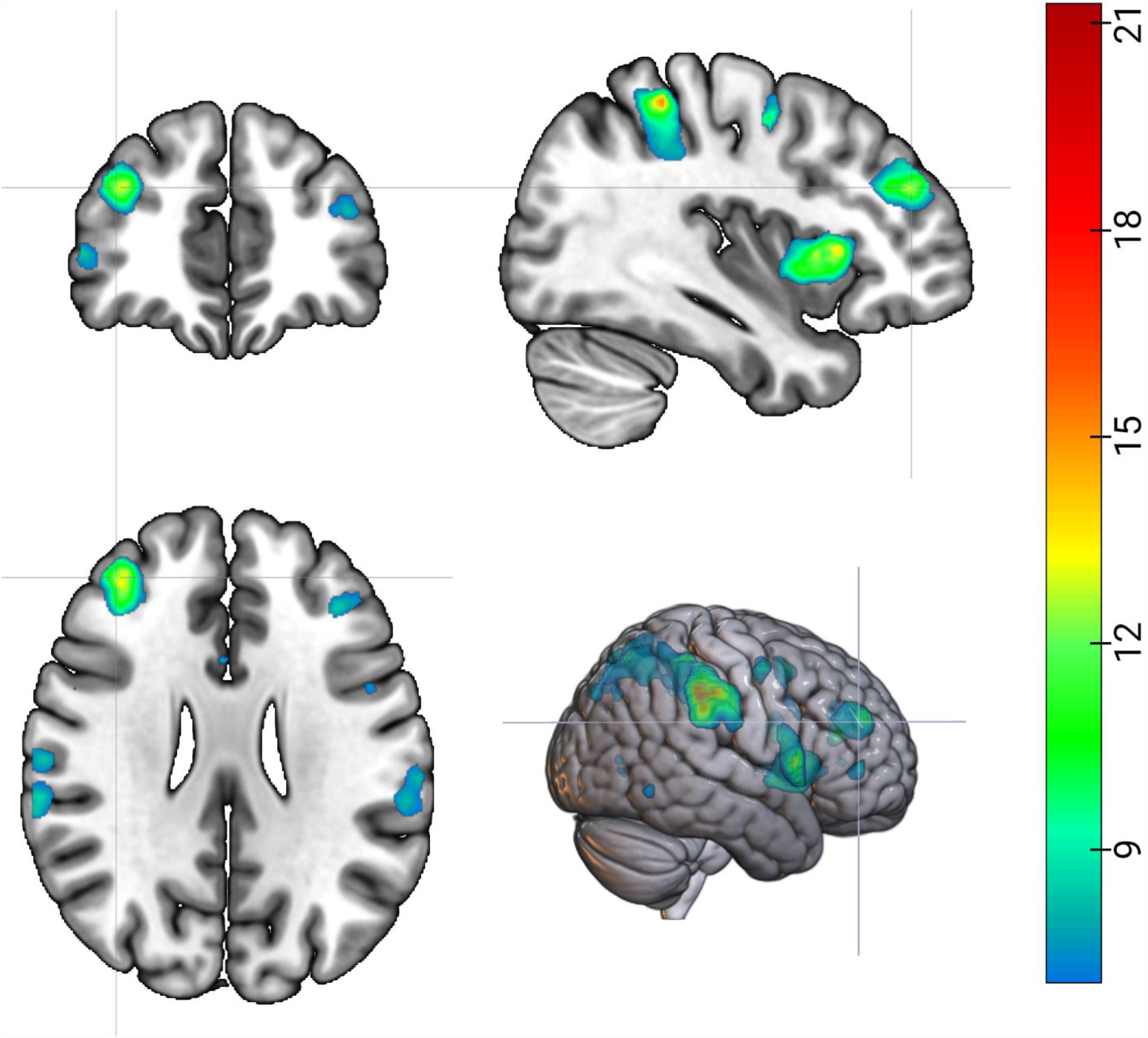
Significantly anticorrelated clusters in both left and right dorsolateral prefrontal cortex were observed at group level corresponding to the anticipated stimulation sites in MNI space. p_FWE,Peak_ < 0.05, height threshold T = 7.07; Color bar: T = [7.07, 21.28]; Left is right

Results further showed that the centroid of the individual most anticorrelated cluster in the DLPFC (i.e., the “ideal”, functionally defined stimulation location, [13]) was within a Euclidean distance of 13 mm of both actual stimulation targets (left: 11.9 ± 5 mm; right: 12.3 ± 5.9 mm (Mean ± SD)). When using stimulation targets as seed to determine voxel-wise functional connectivity to the whole-brain, we observed a significant positive correlation between left and right DLPFC seeds and clusters in bilateral AI, anterior cingulate, supplementary motor area, as well as other frontal and parietal regions (see Table 2). Significant negative correlations were observed for several temporal clusters as well as frontal and occipital regions (see Table 2). There was no correlation between any of the stimulation target connectivities and symptom scores at baseline (p ≥ 0.05).

**Table 2:** Functional connectivity before and after treatment. Only clusters with p_FWE-corr_ ≤ 0.05, following p_uncorr_ = 0.001 are displayed

| Measurement, Seed | Region | MNI<sub>Peak</sub> |  |  | P<sub>FWE-corr</sub> |  |
| --- | --- | --- | --- | --- | --- | --- |
|  | AAL | X | Y | Z | cluster | peak |
| **M1: Baseline rs-fMRI (n = 20)** |  |  |  |  |  |  |
| **DLPFC L** |  |  |  |  |  |  |
| correlation | AI L | -54 | 12 | 2 | <0.001 | <0.001 |
|  | Supp. Motor Area R | 4 | 20 | 48 | <0.001 | 0.079 |
|  | Frontal Mid L | -40 | 36 | 30 | <0.001 | 0.127 |
|  | Frontal Inf Tri R | 38 | 34 | 26 | <0.001 | 0.317 |
|  | Frontal Sup L | -18 | 10 | 68 | 0.012 | 0.179 |
|  | Frontal Inf Oper R | 48 | 20 | -2 | 0.021 | 0.189 |
|  | Frontal Sup Orb L | -20 | 54 | -6 | 0.045 | 0.450 |
| anticorrelation | Temporal Inf R | 44 | -62 | -6 | <0.001 | 0.276 |
| **DLPFC R** |  |  |  |  |  |  |
| correlation | Frontal Inf Tri R | 28 | 30 | 28 | <0.001 | 0.007 |
|  | Insula R | 42 | 18 | -6 | <0.001 | 0.011 |
|  | Insula L | -26 | 22 | 6 | <0.001 | 0.030 |
|  | Precuneus R | 10 | -60 | 58 | <0.001 | 0.046 |
|  | Cingulum Mid | 4 | 12 | 36 | <0.001 | 0.048 |
|  | Frontal Mid | -32 | 38 | 32 | <0.001 | 0.108 |
|  | Parietal Inf R | 46 | -40 | 56 | <0.001 | 0.127 |
|  | Frontal Sup R | 18 | 8 | 70 | 0.004 | 0.487 |
| anticorrelation | Frontal Sup L | -14 | 50 | 32 | <0.001 | 0.001 |
|  | Temporal Pole Mid R | 48 | 14 | -38 | <0.001 | 0.033 |
|  | Frontal Mid Orb L | -2 | 64 | -10 | <0.001 | 0.034 |
|  | Temporal Mid L | -54 | -4 | -22 | <0.001 | 0.202 |
|  | Angular L | -54 | -68 | 28 | <0.001 | 0.256 |
|  | Precuneus | 0 | -52 | 22 | <0.001 | 0.334 |
|  | Cingulum Ant R | 4 | 50 | 18 | 0.021 | 0.064 |
| **M2: Post-treatment rs-fMRI: Active TBS (n = 15)** |  |  |  |  |  |  |
| **DLPFC L** |  |  |  |  |  |  |
| correlation | Frontal Mid L | -42 | 46 | 24 | <0.001 | 0.825 |
|  | Frontal Mid L | -32 | 8 | 64 | 0.009 | 0.910 |
| anticorrelation | Occipital Inf R | 26 | -90 | 0 | <0.001 | 0.377 |
| **DLPFC R** |  |  |  |  |  |  |
| correlation | Frontal Sup L | -28 | 34 | 36 | <0.001 | <0.001 |
|  | Frontal Mid R | 30 | 34 | 36 | <0.001 | 0.001 |
|  | Cingulum Ant R | 4 | 30 | 30 | <0.001 | 0.003 |
|  | Precuneus | -6 | -48 | 56 | <0.001 | 0.055 |
|  | Insula L | -28 | 20 | 10 | 0.001 | 0.267 |
|  | Insula R | 30 | 18 | 8 | 0.003 | 0.226 |
|  | Frontal sup R | 18 | 6 | 58 | 0.004 | 0.406 |
|  | Parietal Inf L | -52 | -38 | 48 | 0.007 | 0.248 |
|  | Frontal Inf Orb L | -32 | 44 | -18 | 0.007 | 0.432 |
|  | Supramarginal R | 66 | -40 | 30 | 0.016 | 0.936 |
| anticorrelation | Temporal Mid R | 56 | -8 | -24 | <0.001 | 0.018 |
|  | Temporal Pole Mid R | 46 | 12 | -30 | <0.001 | 0.072 |
|  | Temporal Pole Sup L | -38 | 18 | -22 | <0.001 | 0.149 |
|  | Rectus | 0 | 48 | -28 | <0.001 | 0.675 |
|  | Precuneus L | -10 | -52 | 24 | 0.009 | 0.521 |
| **M2-M1: rs-fMRI changes rs-fMRI: Active TBS (n = 15)** |  |  |  |  |  |  |
| **DLPFC L** |  |  |  |  |  |  |
| FC reduction | Frontal Inf Orb L | -42 | 26 | -2 | <0.001 | 0.005 |
|  | Frontal Inf Orb R | 50 | 20 | -6 | 0.006 | 0.0273 |
| **DLPFC R** |  |  |  |  |  |  |
| no effect | - | - | - | - | - | - |

To validate the hypothesis that sgACC-DLPFC connectivity predicts antidepressant treatment efficacy of stimulation targets [11-14, 16], we next determined whether baseline anticorrelation strength between individual stimulation targets and sgACC could predict antidepressant response in the active stimulation group. This analysis revealed no significant findings (p ≥ 0.05). However, shorter Euclidean distances between the left stimulation target and the functionally defined “ideal” target (in terms of highest anticorrelation with the sgACC) for all subjects were significantly correlated with relative symptom improvement (% reduction in HAMD-17 score; R_Pearson_ = 0.4664; p = 0.0382, next observation carried backwards (NOCB) for missing values at two weeks post treatment). For post-treatment depression scores in the active stimulation group, we found correlations with left DLPFC-sgACC FC: R_Pearson_ = 0.5603; p = 0.0581 (n = 12 available at two weeks post treatment) and R_Pearson_ = 0.6148; p = 0.0147 (n = 15, NOCB).

Moreover, we observed a significant correlation (R_Pearson_ = 0.6148; p = 0.0147) between the absolute individual HAMD-17 reduction and the anticorrelation between sgACC and left individual stimulation targets when connectivity values were extracted from a normative connectivity map taken from [13], which is based on 2000 twenty-eight– minute resting-state scans from 1000 participants of the Human Connectome Project, (see Fig 2).

**Figure 2.**
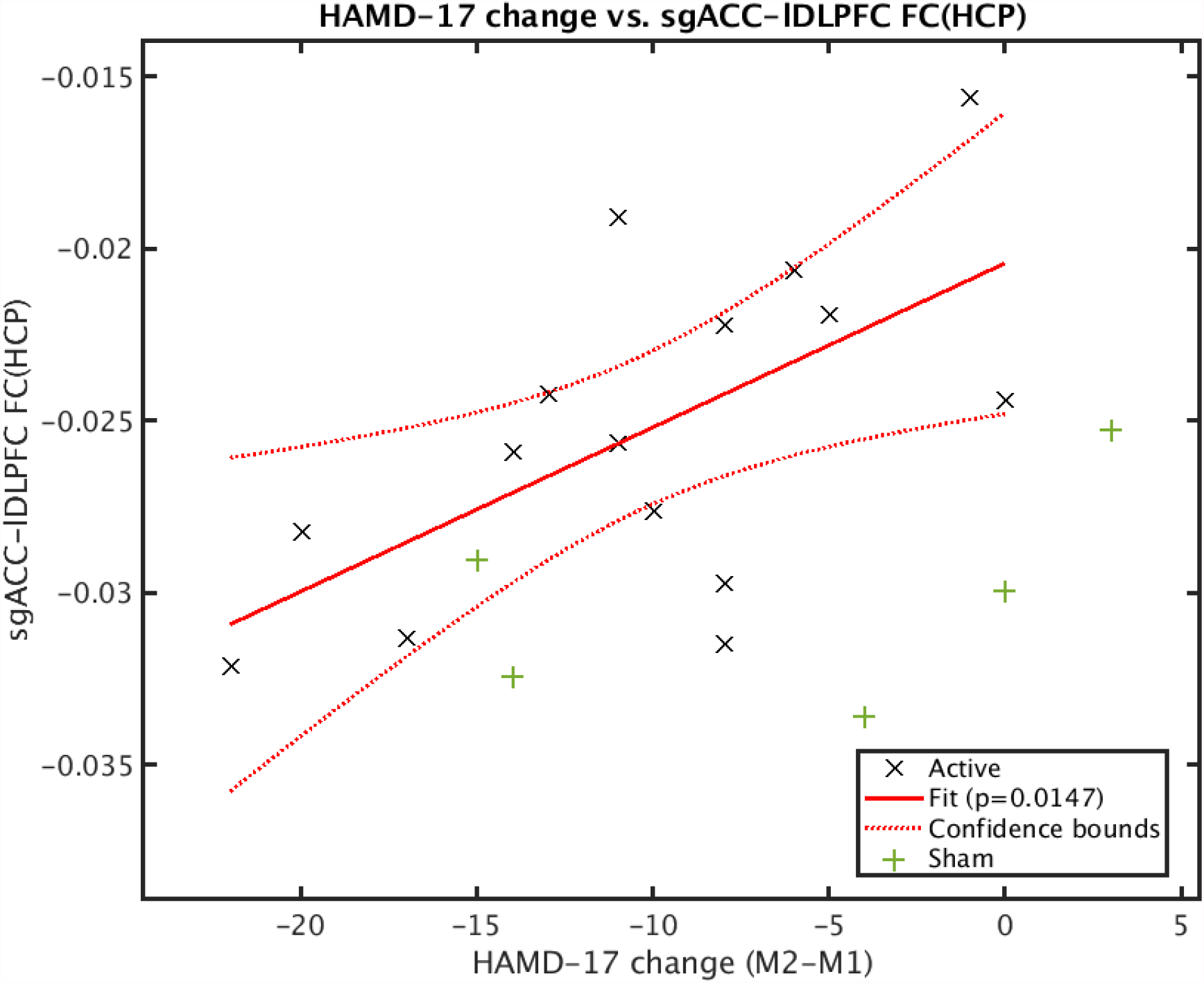
There was a significant correlation between reduction in depressive symptoms in the verum group, as monitored with HAMD-17 scores, and functional connectivity towards the sgACC of the stimulation site, extracted from a FC map derived from the Human Connectome Project data.

### Functional connectivity change over time

We first probed functional connectivity changes in the stimulation group. When placing the seed in the left and right stimulation target, respectively, there was no significant change for stimulation target-to-sgACC connectivity. However, we observed a significant reduction in functional connectivity between the left stimulation target and bilateral AI (see Fig 3). No significant connectivity changes were observed for the right stimulation target. For the sham stimulation group, no significant connectivity changes were observed. Moreover, there was no significant correlation between connectivity changes and a change in any clinical symptom scale.

**Figure 3.**
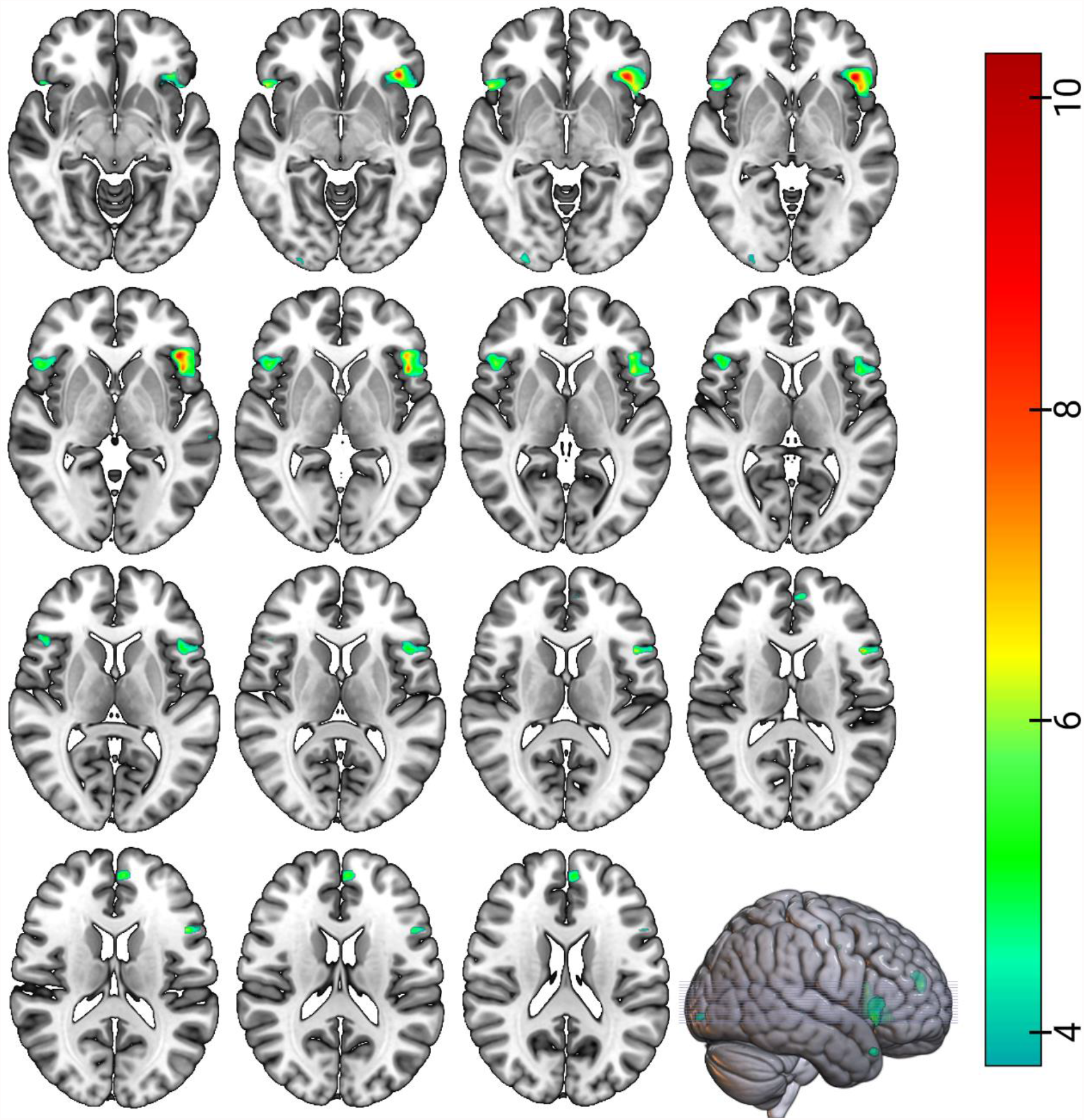
In the bilateral anterior insula regions we observed significant (p_uncorr_ < 0.001, p_FWE Cluster_ < 0.05) functional connectivity (FC) reductions following three weeks of active theta-burst stimulation-treatment. FC-reduction in the superior frontal gyrus was significant only at p_uncorr_ < 0.001. Height threshold T = 3.79 (p_uncorr_ <0.001); Color bar: T = [3.79, 10.28]; Left is right

### Functional connectivity after treatment

When determining if post-treatment connectivities (see Table 2) correlate with post-treatment symptom scores, we observed a significant positive correlation (see Figure 4), between left stimulation target-sgACC connectivity and residual HAMD-17 scores (R_Pearson_ = 0.587, p = 0.045) as well as IDS-C (R_Pearson_ = 0.675, p = 0.016) and a trend towards significance for BDI-II (R_Pearson_ = 0.558, p = 0.060). That is, the higher residual symptom load, the less anticorrelated was the left stimulation target with the sgACC.

**Figure 4.**
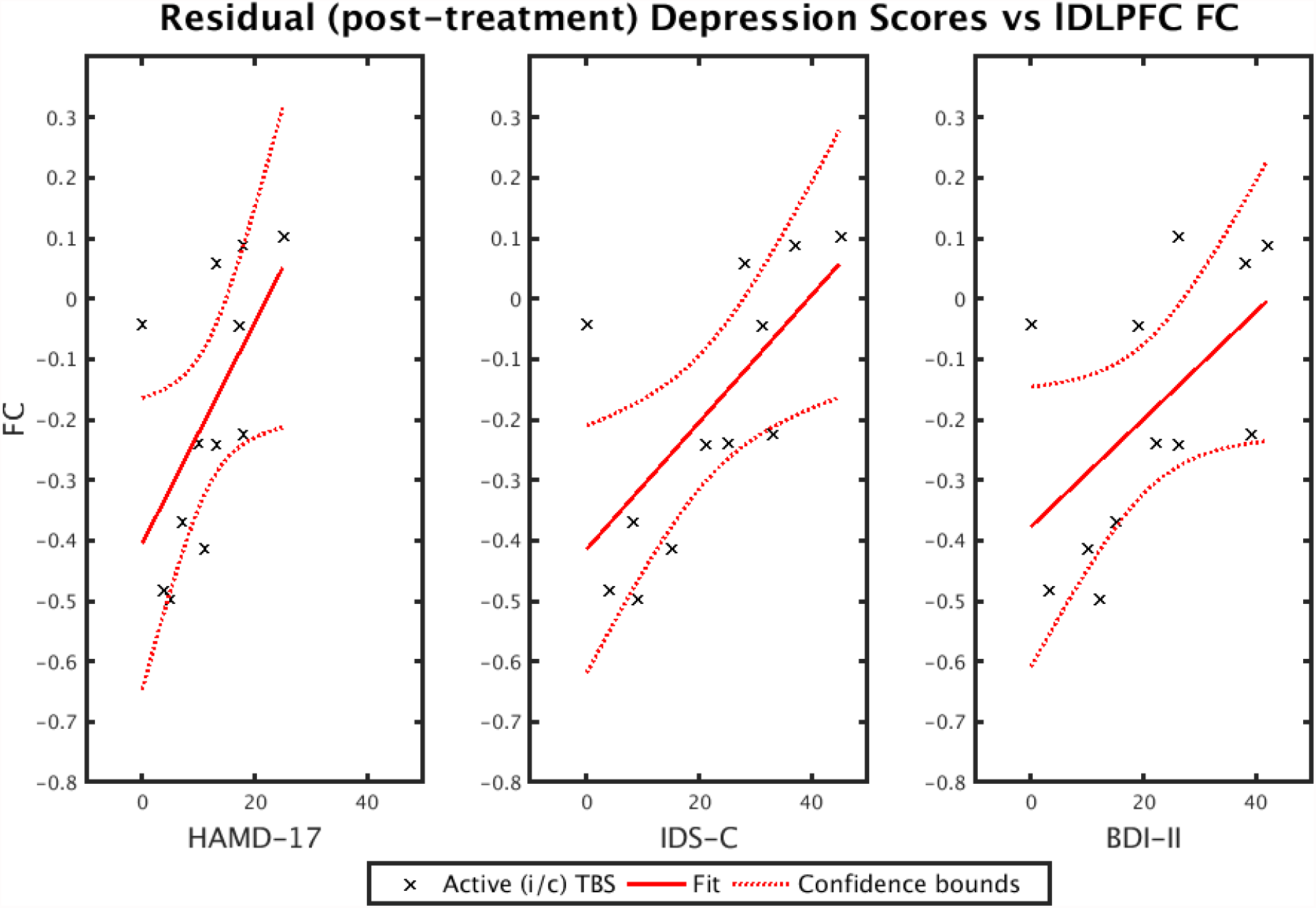
Depression scores after three weeks of active theta-burst stimulation were correlated with post-treatment functional connectivity of the left dorsolateral prefrontal cortex stimulation site: HAMD-17: R_Pearson_ = 0.587, p = 0.045; IDS-C: R_Pearson_ = 0.675, p = 0.016; BDI-II: R_Pearson_ = 0.558, p = 0.060; Please note, that due to incomplete data collection, depression scores two weeks post treatment were not fully available for each of the 15 treated subjects, thus n = 12.

## Discussion

This study assessed the effects of a three-week bilateral sequential theta-burst stimulation, comprising left iTBS and right cTBS, on functional connectivity in treatment-resistant depression. The results were based on data from 20 patients of a prematurely closed trial, and showed clinical improvement of TRD over both groups (active and sham) but no significant interaction effect of group, or group and time. The effect in the active TBS group was larger though, than in the combined analysis. In the active treatment group (n = 15), 53% (8) of the patients responded (including remitters) and 20% (3) remitted, while in the sham treatment group (n = 5), 40% (2) responded and 0% (0) remitted. The acquired resting-state fMRI data showed that the stimulation targets were anticorrelated with the sgACC in all patients, and that this association was important for treatment response. Shorter distances between actual stimulation targets, and ideal ones defined by anticorrelation with the sgACC according to [13] improved treatment response. Following active TBS treatment, no changes in stimulation target-to-sgACC connectivity, but a reduction of FC between the left stimulation target and bilateral anterior insula was found.

Regarding the efficacy of the stimulation protocol in the unique TRD patient collective, our data show improvement of depression in both the active and sham TBS group. While there is considerable evidence for the efficacy of various rTMS protocols in major depression, there is little data available for new and time efficient TBS protocols, which is also why there are no firm recommendations yet [1]. Accordingly, the herein applied cTBS right and iTBS left protocol has been rated to have a probable but not yet fully proven antidepressant effect [1]. While our response rate of 53% is roughly comparable to previous well-powered trials investigating unilateral iTBS of the left DLPFC [3, 35], we could not find a similarly high response rate for bilateral TBS as for example Li et al., who observed almost 67% [2]. Our study would therefore rather suggest an efficacy comparable to unilateral iTBS. In line with our high response rate to sham stimulation (40%), Li et al. also observed a considerable sham effect, especially for a subgroup with a low level of refractoriness [2], a trait not assessed in our sample. It is conceivable, that other factors, such as regular social interaction to health-care personnel or having three-week daily routine may have contributed to the antidepressant effect. However, it should be kept in mind that due to the premature termination of our trial, our sham group is particularly small (n=5), thus limiting transferability and the informative value of our results. Moreover, the response rate observed in both groups could also reflect the heterogeneity of the unique TRD patient population, where individual aspects ranging from biological, psychological, and sociocultural factors are likely to contribute to treatment-resistance [36]. Nonetheless, it should be noted that the remission rate of the comparable TRD patient collective in the third treatment step of the important STAR^*^D study was only 13.7% [37]. Therefore, our remission rate of 20% following TBS, as well as the larger effect size in the active treatment group may be an argument in favor for the antidepressant effect of bilateral theta-burst stimulation. Likewise, the reductions of depression scores were markedly higher in the verum group as in the sham group. However, the direct comparison cannot be made with certainty, due to the skewed groups along with limited size of the sham group in our study.

Despite the limitations our study has in determining the effectiveness of the specific protocol, our neuroimaging data may contribute to the biological understanding of TRD, and the role bilateral TBS could have in its modulation. Most importantly, our study provided further evidence of the importance of an anticorrelation between stimulation targets primarily in the left DLPFC for antidepressant treatment success. First, stimulation targets in the left and right DLPFC were located within clusters that showed significant anticorrelation with the sgACC. Second, the distance of the left target to the largest baseline DLPFC-to-sgACC anticorrelation showed a significantly positive correlation with treatment response. This result partly confirms previous reports [13]. Third, there was a significant correlation between treatment response and the FC between sgACC and left individual stimulation targets when connectivity values were extracted from a normative connectivity map taken from data of the Human Connectome Project. Fourth, after treatment, residual symptom load was significantly correlated with individual left stimulation target-to-sgACC connectivity. The higher residual symptom load, the less anticorrelated the left stimulation target was with the sgACC.

Our study indicates clinical relevance of sgACC-DLPFC connectivity primarily for the left but not right hemisphere. Interestingly, an early PET study using _15_O-water demonstrated a link between mood changes and reciprocal changes in regional blood flow (measured with of sgACC and the right DLPFC. That is, with increasing scores for sadness, blood flow increases in sgACC were accompanied with blood flow decreases in right DLPFC, whereas depression recovery was associated with the reverse pattern of blood flow changes [38]. This is in contradiction to the notion that stimulation of the right DLPFC should be inhibitory in order to counteract its presumed hyperactivity in MDD. Indeed, left HF rTMS and right LF rTMS have contrasting effects on metabolic activity in connected brain areas [39-41]. Despite discrepant findings, our study does not preclude a role of right DLPFC in the antidepressant effect of therapeutic brain stimulation. More studies are needed to reveal the mechanism of action of bilateral sequential DLPFC stimulation involving left excitatory and right inhibitory stimulation.

According to the cognitive theory of depression posing prefrontal control over limbic hyper-activation, one would also expect a change in target-to-sgACC functional connectivity. Specifically, an increase in anticorrelation could be assumed and that such an increase correlates with treatment response. However, we did not observe such effect in our data, corroborating a previous study with a similar sample size, which also does not support this hypothesis [22]. Studies showing an association between sgACC-to-DLPFC connectivity on the individual or group level tend to have twice our sample size [12, 14], thus the absence of such a relationship is most likely related to the insufficient sample size. Considering these aspects, rTMS may therefore not affect cognitive control networks directly, but rather modulate other networks including the DMN, as observed previously [22].

A central new finding in our study is that bilateral TBS leads to a reduction in FC between the left stimulation target and bilateral AI. The AI is part of the SN, the main target network for rTMS in depression and addiction [19] and structural abnormities therein have been shown to be a common neural substrate of psychiatric disorders [42]. A previous study in healthy participants observed an increased target-AI FC when stimulating the left DLPFC with cTBS [21]. Our results showing a reduction of FC upon iTBS to the left (and cTBS to the right) DLPFC are therefore compatible with this observation. However, a treatment-induced FC reduction as observed in our study may be interpreted as a decoupling of left DLPFC with the AI, which would contradict the notion of a strengthened SN upon brain stimulation. In any case, interpretations need to be done with caution, given that we observed no direct correlation with symptom improvement in our data.

Prefrontal and cognitive models of depression posit an insufficient top-down control of the prefrontal cortex over limbic hyperactivity in MDD [18, 43] and non-invasive brain stimulation including rTMS supposedly counteracts this deficit [44]. Our study contributes to these theories by demonstrating modulations within the SN upon bilateral sequential TBS.

### Limitations

Our study has several limitations that compromise the interpretation of its results: First, due to the premature termination, the unintended small sample size, especially in the control group, suggests that interpretations should be made with caution. We also did not correct for multiplicity of whole-brain models and correlations, rendering our study exploratory. Second, accumulating evidence points to the importance of prolonged treatment regimens lasting at least 4 to 6 weeks. Our treatment duration of only 3 weeks may have been too short to determine reliable treatment effects. Third, positive psychological effects of the daily interaction with health-care personnel cannot be ruled out and could also explain improved symptoms without active TBS. Fourth, although patients were resistant to pharmacological treatment and on stable medication, effects and interactions with TBS cannot be ruled out. Finally, although unlikely, a subtle effect of the magnetic field in the sham orientation of the coil may also play a role in the response rates to the sham stimulation.

## Conclusions

Our study investigated the influence of bilateral sequential theta-burst stimulation on functional connectivity in treatment-resistant depression. Imaging results are compatible with previous findings, highlighting the clinical importance of the connection between the dorsolateral prefrontal and the subgenual cingulate cortex. We further show changes within the salience network, a network thought to be crucial for the therapeutic effect of repetitive transcranial magnetic stimulation. Our study thus contributes to the growing body of literature on the effects of transcranial magnetic stimulation on functional connectivity in depression, yet inferences from our limited data about the efficacy of the protocol should be drawn with caution.

## Data Availability

Due to data protection laws, processed, anonymized data is only available from the authors upon reasonable request.

## Author contributions

**Peter Stöhrmann:** Methodology, Data Curation, Processing & Analysis, Writing-Original Draft, Writing-Review & Editing, Visualization; **Godber M Godbersen:** Investigation, Data Acquisition & Curation, Writing-Review & Editing; **Murray B Reed:** Resources, Investigation, Methodology, Data Acquisition & Curation, Writing-Review & Editing; **Jakob Unterholzner:** Investigation, Data Acquisition, Writing-Review & Editing; **Manfred Klöbl:** Methodology, Resources, Investigation, Data Acquisition & Curation, Writing-Review & Editing; **Pia Baldinger-Melich:** Investigation, Data Acquisition, Writing-Review & Editing; **Thomas Vanicek:** Investigation, Data Acquisition, Writing-Review & Editing; **Andreas Hahn:** Conceptualization, Methodology, Investigation, Data Acquisition & Curation, Writing-Review & Editing; **Rupert Lanzenberger:** Funding, Conceptualization, Methodology, Resources, Writing-Review & Editing, Supervision, Project administration; **Siegfried Kasper:** Funding, Supervision, Conceptualization, Resources, Project administration, Writing-Review & Editing; **Georg S. Kranz:** Conceptualization, Methodology, Writing-Original Draft, Writing-Review & Editing, Supervision, Project administration.

## Acknowledgements

This research was funded in whole, or in part, by the Austrian Science Fund (FWF) [Grant number KLI 551, PI: S. Kasper]. For the purpose of open access, the author has applied a CC BY public copyright license to any Author Accepted Manuscript version arising from this submission. M.B. Reed is a recipient of a DOC fellowship of the Austrian Academy of Sciences at the Department of Psychiatry and Psychotherapy, Medical University of Vienna. We would like to express our gratitude towards Robin Cash, Andrew Zalesky, Luca Cocchi and their colleagues for providing us with FC-maps derived from the Human Brain Project and valuable insights, as well as Jonathan Downar for his assistance in the conceptualization of the study. We would further like to thank Richard Frey, Gregor Gryglewski, Marius Hienert, Marie Spies, Christoph Kraus, Alexander Kautzky, Arkadiusz Komorowski, Paul Michenthaler for clinical support, and Sebastian Ganger for technical support, and all additional staff and students from the Neuroimaging Lab (NIL) involved in the realization of this research.

## Data availability statement

Please contact with any questions or enquiries.

